# Chronic conditions and risk of severe outcomes among adults aged 18-59 years with medically-attended RSV illness in the United States

**DOI:** 10.1101/2025.06.17.25329736

**Authors:** Daniel K. Nomah, Linwei Li, John Shen, Neloufar Rahai, Meng Wang, Isabella Lelis, Miao Jiang, Genelle Goodhue, Beverly Francis, Sophia Monaghan, Claire Newbern, Catherine A. Panozzo, Elissa Wilker, Zhe Zheng

## Abstract

**Background:** Respiratory Syncytial Virus (RSV) can cause severe respiratory disease. While risks among older adults are well established, less is known about RSV severity in younger adults with chronic conditions.

**Methods:** A retrospective cohort study using U.S. commercial and Medicaid claims data was conducted to evaluate outcomes in adults aged 18–59 with medically attended RSV (MA-RSV) in 2022-24. Chronic conditions were assessed during a 12-month baseline period prior to the MA-RSV. Severe outcomes, including RSV-associated hospitalization, RSV-associated intensive care unit admission, mechanical ventilation, and death, were assessed within 30 days following RSV diagnosis. Adjusted risk ratios (aRRs) and 95% confidence intervals (CIs) were estimated using multivariable models.

**Results:** In both commercially insured (n=1435 in 2022-23; n=1890 in 2023-24) and Medicaid-insured (n=2065) adults with MA-RSV, approximately 75% had ≥1 chronic condition, and over 25% had four or more. Severe RSV outcomes occurred in ∼21% of individuals. The risk of severe outcomes substantially increased with the number of chronic conditions. Among commercially insured adults, the aRR for those with two to three chronic conditions compared to none was 10.06 (95% CI: 4.80–21.08) in the 2022–2023 and 13.74 (95% CI: 5.95–31.73) in 2023–2024. For those with four or more conditions, the aRRs were 21.39 (95% CI: 10.26–44.60) and 38.03 (95% CI: 16.61–87.11), respectively. In the Medicaid cohort (2023–2024), the aRR was 7.68 (95% CI: 4.63–12.76) for individuals with two to three conditions and 13.85 (95% CI: 8.36– 22.95) for those with four or more.

**Conclusion:** Adults aged 18–59 years with chronic conditions are at increased risk for severe RSV illness, supporting broader prevention strategies, including vaccination.

## Introduction

Respiratory syncytial virus (RSV) is a major cause of morbidity and mortality in pediatric and elderly populations; however, adults aged 18–59 years with chronic conditions may also be at elevated risk [1–4]. Recently, the U.S. Advisory Committee on Immunization Practices (ACIP) recommended RSV vaccination for adults aged 50–59 years with certain chronic conditions, including cardiovascular disease, chronic respiratory disease, diabetes, and immunodeficiency, who have an elevated risk of severe outcomes related to RSV [5]. Many of these conditions are established risk factors for severe outcomes from RSV in older adults and other respiratory illnesses for adults of all ages [6,7,8], contributing to greater healthcare utilization, including longer hospital stays and intensive care admissions [9,10].

While the CDC has identified chronic conditions that elevate the risk of severe RSV outcomes in adults aged 50 years and older, limited data exist to inform which adults aged 18–49 years are at greatest risk for severe RSV disease. This study aimed to assess the prevalence of chronic conditions that may increase the risk of severe RSV outcomes among adults aged 18–59 years diagnosed with RSV and to evaluate the association between those chronic conditions and severe RSV outcomes, using large U.S. administrative datasets to inform public health strategies and optimize prevention efforts in this younger high-risk population.

## Methods

### Data Sources

This study used deidentified administrative healthcare claims from two large U.S. insurance databases: Optum’s Clinformatics® Data Mart (CDM) and Merative™ MarketScan® Multi-State Medicaid (MDCD). The CDM includes data from millions of individuals nationwide enrolled in commercial or Medicare Advantage plans, while the MDCD database captures Medicaid enrollees.

All data were fully deidentified and compliant with the Health Insurance Portability and Accountability Act (HIPAA) requirements [11]. As this analysis involved secondary data without patient identifiers or direct contact, institutional review board (IRB) approval and informed consent were not required [12]. The study followed the Strengthening the Reporting of Observational Studies in Epidemiology (STROBE) guidelines for observational research [13].

### Study Design and Population

In this retrospective cohort study of adults aged 18–59 years with medically-attended RSV (MA-RSV), we assessed the prevalence of chronic conditions, RSV-related healthcare resource utilization (HCRU), and the association between chronic conditions and severe RSV outcomes. The study included two respiratory disease seasons for the commercially-insured cohorts (July 1, 2022 to June 30, 2023, and July 1, 2023 to June 30, 2024), and a single season for the Medicaid-insured cohort (July 1, 2022 to June 30, 2023).

Eligible individuals were required to have at least one MA-RSV encounter (outpatient, emergency department, or inpatient) during the study period. The index date was defined as the first MA-RSV encounter observed in each RSV season. MA-RSV was identified using International Classification of Diseases, Tenth Revision, Clinical Modification (ICD-10-CM) codes for MA-RSV in conjunction with lower respiratory tract infection (LRTI) codes **(Supplementary Table S1)**, with both RSV and LRTI codes allowed in any diagnostic position [14]. Only the first observed MA-RSV episode per season was considered to avoid duplicate individuals with multiple events in a single respiratory season. Individuals were required to have continuous health plan enrollment for at least 365 days prior to the MA-RSV index date (allowing for a single ≤30-day gap).

Individuals who were not between 18 and 59 years old at the time of their MA-RSV index date or had missing key demographic data (age, sex) were excluded; patient attrition is detailed in **Supplementary Table S2.** Analyses were limited to participants without any documented RSV vaccination during the baseline or follow-up period, as RSV vaccines were not widely available for this age group, and to ensure that analyses of severe outcomes reflected RSV-related morbidity among unvaccinated individuals. A visual summary of the study design, including cohort entry and follow-up windows, is provided in the supplementary materials **(Supplementary Figure S1)**.

### Measures

Chronic conditions were assessed using ICD-10-CM diagnosis and procedure codes during the 365-day baseline period prior to the MA-RSV index date. Conditions were based on the definitions used in analyses of the U.S. CDC RSV-NET hospitalization surveillance system [15], which identifies common risk factors for severe RSV. These included cardiovascular disease (e.g, coronary artery disease [CAD] and heart failure), obesity, diabetes, chronic respiratory disease (e.g., chronic obstructive pulmonary disease [COPD] and asthma), chronic kidney disease, chronic liver disease, immunocompromised status, neurologic conditions, and blood disorders. Full definitions and code lists, which were based on and modified from the RSV vaccine effectiveness study by Payne et al. [16], are provided in **Supplementary Table S3**. RSV-associated hospitalizations or ICU admissions were defined as hospitalizations or ICU admissions with an in any position. All-cause mortality was assessed only in the commercially insured cohorts, as death data were unavailable in the Medicaid cohort. Other healthcare utilization outcomes of interest included antibiotic use, oxygen support, and length of stay for hospitalizations and ICU admissions. Measure definitions are detailed in **Supplementary Table S4**. Individuals were followed from the MA-RSV index date to 30 days post-index, death, or hospitalization discharge, whichever occurred first.

### Statistical Analysis

Patient characteristics, chronic conditions, and HCRU were described using summary statistics. Categorical variables were reported as counts and percentages, while continuous variables were summarized using means and standard deviations and/or medians and interquartile ranges. In assessing the prevalence of chronic conditions among individuals diagnosed with RSV in the commercially-insured cohort, data were pooled across the 2022–2023 and 2023–2024 seasons. Associations between chronic conditions and severe RSV outcomes were estimated using multivariable generalized linear models with a Poisson distribution, log link function, and robust standard errors [17,18]. The severe RSV outcome was defined as a composite of RSV-associated hospitalization, ICU admission, mechanical ventilation (MV), or all-cause mortality within 30 days of the MA-RSV index date. Adjusted risk ratios (aRRs) with 95% confidence intervals (CIs) were reported, adjusting for age and sex. Comorbidities were evaluated in two ways: (1) by the number of chronic conditions (categorized as 0, 1, 2–3, or ≥4 conditions) and (2) by individual chronic conditions, included simultaneously in the model to assess their independent associations with severe RSV outcomes.

Sensitivity analyses were conducted to assess the robustness of findings. First, analyses restricted to encounters with RSV coded in the primary diagnosis position were performed to address potential misclassification. Second, narrower definitions of chronic conditions were applied, focusing on more severe disease forms based on CDC criteria [19] (definitions provided in **Supplementary Table S5**). Finally, effect modification by age was explored by evaluating the association between chronic conditions and severe outcomes across different age subgroups, additionally adjusting for sex.

All analyses were conducted separately for the commercially insured and Medicaid-insured populations using the Substantiate application of the Aetion Evidence Platform® (AEP) version 5.3.3, with supplemental analyses performed in R statistical software (version 4.4.0; R Foundation for Statistical Computing) using the ‘*sandwich*’ and ‘*lmtes*t’ packages.

## Results

### Baseline Characteristics

In the commercially-insured cohort, 1,435 adults aged 18–59 years were identified with medically attended RSV (MA-RSV) during the 2022–2023 season (median age 47.0 years [IQR: 35.0–55.0]; 64% female) and 1,890 during the 2023–2024 season (median age 48.0 years [IQR: 37.0–55.0]; 60% female). In the Medicaid-insured cohort, 2,065 adults aged 18–59 years were identified during the 2022–2023 season (median age 38.0 years [IQR: 27.0–50.0]; 72% female).

Compared with commercially-insured adults, Medicaid-insured individuals were younger and more frequently diagnosed with RSV in emergency departments or inpatient settings (Table 1).

**Table 1:** Baseline Demographic and Clinical Characteristics of Adults aged 18-59 with Medically Attended RSV in the U.S., 2022–24.

| Characteristic | Commercially-Insured<br>Cohort<br>(2022–23)<br>N=1,435 | Commercially-Insured<br>Cohort<br>(2023–24)<br>N=1,890 | Medicaid-Insured<br>Cohort<br>(2022–23)<br>N=2,065 |
| --- | --- | --- | --- |
| **Age, in years** |  |  |  |
| Mean (SD) | 44.23 (11.89) | 45.28 (11.65) | 38.52 (12.80) |
| Median [IQR] | 47.00 [35.00, 55.00] | 48.00 [37.00, 55.00] | 38.00 [27.00, 50.00] |
| **Age groups, in years, n (%)** |  |  |  |
| 18–29 | 208 (14.5%) | 234 (12.4%) | 609 (29.5%) |
| 30–39 | 279 (19.4%) | 317 (16.8%) | 490 (23.7%) |
| 40–49 | 314 (21.9%) | 447 (23.7%) | 423 (20.5%) |
| 50–59 | 634 (44.2%) | 892 (47.2%) | 543 (26.3%) |
| **Gender, n (%)** |  |  |  |
| Female | 912 (63.6%) | 1,142 (60.4%) | 1,496 (72.4%) |
| Male | 523 (36.4%) | 748 (39.6%) | 569 (27.6%) |
| **Race/Ethnicity, n (%)<sup>1</sup>** |  |  |  |
| White | 687 (47.9%) | 872 (46.1%) | 1,192 (57.7%) |
| Black | 154 (10.7%) | 217 (11.5%) | 439 (21.3%) |
| Hispanic | N/A | N/A | 216 (10.5%) |
| Asian | 45 (3.1%) | 41 (2.2%) | N/A |
| Other/Unknown | 67 (4.7%) | 72 (3.8%) | 102 (4.9%) |
| Missing | 482 (33.6%) | 688 (36.4%) | 116 (5.6%) |
| **Type of Healthcare Encounter at RSV**<br>**Diagnosis** |  |  |  |
| Outpatient visit | 718 (50.0%) | 960 (50.8%) | 651 (31.5%) |
| ED visit | 446 (31.1%) | 554 (29.3%) | 971 (47.0%) |
| Hospitalization | 271 (18.9%) | 376 (19.9%) | 443 (21.5%) |
| **Underlying Chronic Conditions** |  |  |  |
| Any condition of interest (broad definition)<sup>2</sup> | 1,062 (74.0%) | 1,453 (76.9%) | 1,573 (76.2%) |
| Any condition of interest (narrow definition)<sup>3</sup> | 861 (60.0%) | 1,190 (63.0%) | 1,308 (63.3%) |

Abbreviations - MA-RSV: Medically attended Respiratory Syncytial Virus; SD: Standard Deviation; IQR: Interquartile Range; ED visit:
Emergency Department visit.
<sup>1</sup> Hispanic ethnicity is no longer reported in Optum CDM (as of v0.9). Race in Optum CDM is in the process of changing from third-party imputed to a self-reported source.
<sup>2</sup> Broad definition of high risk for severe RSV, defined as diagnosis with one or more of the following underlying medical conditions in medical claims: Cardiovascular disease (e.g. heart failure, coronary artery disease, congenital heart disease), Obesity, Diabetes, Chronic kidney disease, Neurologic condition, Immunocompromised conditions (e.g. rheumatologic/autoimmune/inflammatory disease, hematological malignancy, solid malignancy, transplant, other intrinsic immune condition or immunodeficiency), Chronic respiratory disease (e.g. COPD, Asthma, interstitial lung disease, cystic fibrosis, and emphysema), Chronic liver disease, Blood disorders
<sup>3</sup> Narrow definition of high risk for severe RSV, defined as diagnosis with one of the following underlying medical conditions in medical claims: Chronic cardiovascular disease (e.g., heart failure, coronary artery disease, or congestive heart failure [excluding isolated hypertension]), Chronic lung or respiratory disease (e.g., COPD, emphysema, asthma, interstitial lung disease, or cystic fibrosis), End-stage renal disease or dependence on hemodialysis or other renal replacement therapy, Diabetes mellitus complicated by chronic kidney disease, neuropathy, retinopathy, or other end-organ damage, or requiring treatment with insulin or sodium-glucose cotransporter-2 (SGLT2) inhibitor, Neurological or neuromuscular conditions causing impaired airway clearance or respiratory muscle weakness (e.g., poststroke dysphagia, amyotrophic lateral sclerosis, or muscular dystrophy [excluding history of stroke without impaired airway clearance]), Chronic liver disease (e.g., cirrhosis), Chronic hematologic conditions (e.g., sickle cell disease or thalassemia), Severe obesity (body mass index $\geq 40$ kg/m<sup>2</sup>), Moderate or severe immune compromise (e.g. active treatment for malignancies, SOT or islet transplant, hematopoietic cell transplant, primary immunodeficiency, autoimmune disease (e.g. lupus, psoriasis, multiple sclerosis))

### Prevalence of Chronic Conditions

The prevalence of chronic conditions among individuals with MA-RSV is presented in **Figure 1**. The majority of adults with MA-RSV had at least one chronic condition: 75.6% among commercially insured individuals and 76.2% among Medicaid-insured individuals.

**Figure 1:**
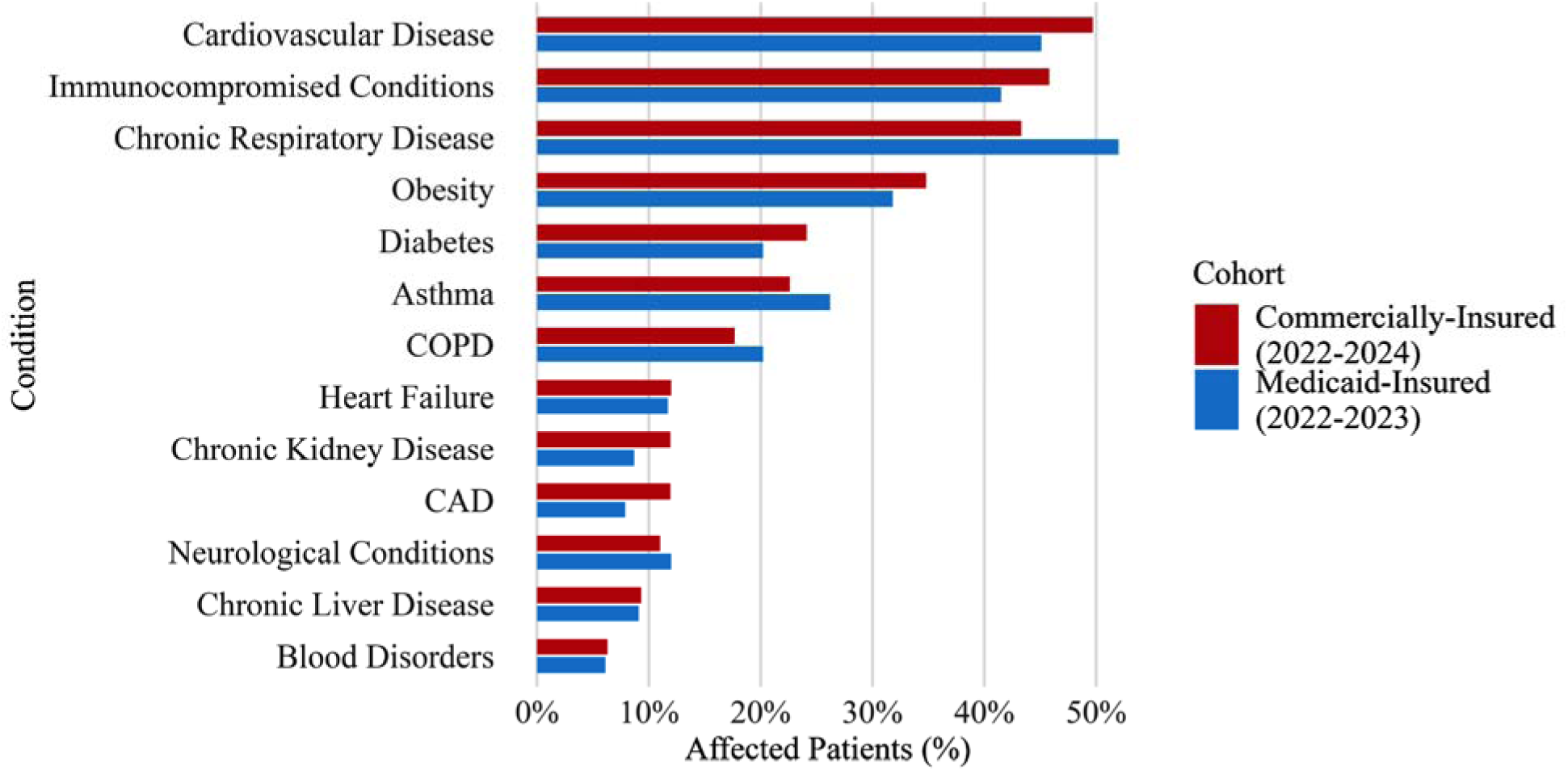
Prevalence of Underlying Chronic Conditions Among Adults Aged 18-59 with Medically Attended RSV in Commercially- (2022–2024) and Medicaid-Insured (2022–2023) Abbreviations: COPD: Chronic Obstructive Pulmonary Disease; CAD: Coronary Artery Disease; MA-RSV: Medically Attended Respiratory Syncytial Virus. Footnote: Cardiovascular disease includes coronary artery disease (CAD) and heart failure (HF); chronic respiratory disease includes chronic obstructive pulmonary disease (COPD) and asthma.

Multimorbidity was common with over 55% of individuals having two or more conditions and more than 25% in each cohort had four or more chronic conditions. The most frequently observed conditions across both datasets were cardiovascular disease, chronic respiratory disease, immunocompromised status, obesity, and diabetes.

### Healthcare Resource Utilization and Severe Outcomes in 30-day Post-Index

Among commercially insured adults aged 18-59 years with MA-RSV, 20.9% were hospitalized in the 2022-23 season. Within 30 days of RSV diagnosis, 9.2% were admitted to the ICU, 4.3% required MV, 30.4% received antibiotics, and 12.4% required oxygen support. In the 2023-24 season for the commercially-insured cohort, healthcare resource utilization was similar: 21.8% hospitalized, 9.7% ICU admission, 4.3% MV, 29.2% antibiotic use, and 12.9% oxygen support. In the 2022-2023 Medicaid-insured cohort, 24.5% were hospitalized, 11.0% admitted to the ICU, 4.7% received MV, 31.3% received antibiotics, and 15.5% required oxygen support.

Severe RSV outcomes and HCRU were especially common among individuals with chronic conditions (**Figure 2** and **Supplementary Table S6)**. The occurrence of severe outcomes increased with age. In the commercially-insured cohort, hospitalization occurred in 13.6% of individuals aged 18–29 years, rising to 31.1% among those aged 50–59 years during the 2022–23 season. Similar age-related increases were observed for ICU admission (7.0% to 13.8%), oxygen support (4.0% to 18.4%), and MV (2.0% to 7.4%). These patterns were consistent in the 2023– 24 commercially insured cohort and the 2022–23 Medicaid-insured cohort.

**Figure 2:**
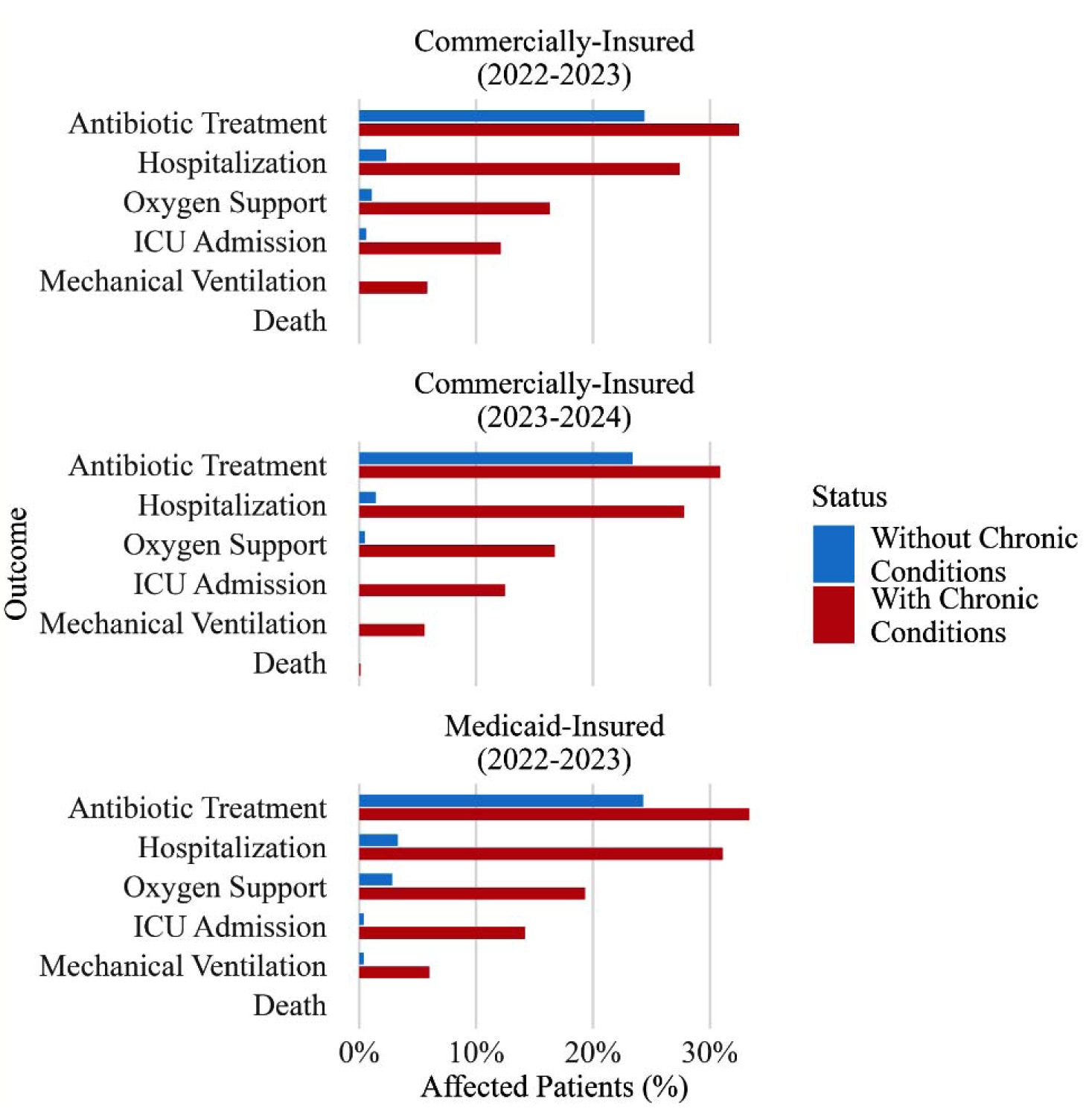
Healthcare Resource Utilization and Severity of Illness Among Adults Aged 18-59 with Medically Attended RSV by Underlying Chronic Condition Status in Commercially- (2022–2024) and Medicaid-Insured (2022–2023) Cohorts. Abbreviations: ICU: Intensive Care Unit; MA-RSV: Medically Attended Respiratory Syncytial Virus. Footnote: All outcomes reported are among individuals with medically attended RSV (MA-RSV) across all care settings (outpatient, emergency department, and inpatient). Oxygen support includes cases where mechanical ventilation (MV) was used.

### Risk Ratio Estimates for Severe Outcomes

Compared to individuals without any chronic conditions, individuals with multiple chronic conditions had a significantly higher risk of severe RSV outcomes in both commercially and Medicaid-insured adults. Among commercially-insured adults, the aRR for those with four or more chronic conditions compared to none was 21.39 (95% CI: 10.26–44.60) in the 2022–23 season and 38.03 (95% CI: 16.61–87.11) in the 2023–24 season. In the Medicaid-insured cohort, the aRR was 13.85 (95% CI: 8.36–22.95). Elevated risk was also observed among individuals with two to three chronic conditions, with aRRs of 10.06 (95% CI: 4.80–21.08) and 13.74 (95% CI: 5.95–31.73) in the commercially insured cohort during 2022–23 and 2023–24 seasons, respectively, and 7.68 (95% CI: 4.63–12.76) in the Medicaid-insured cohort. Even individuals with a single chronic condition had a moderately increased risk, with aRRs of 2.47 (95% CI: 1.03–5.89) and 4.21 (95% CI: 1.67–10.57) in the commercially-insured cohort during 2022–23 and 2023–24 seasons, and 2.66 (95% CI: 1.49–4.76) in the Medicaid-insured cohort (**Figure 3**).

**Figure 3:**
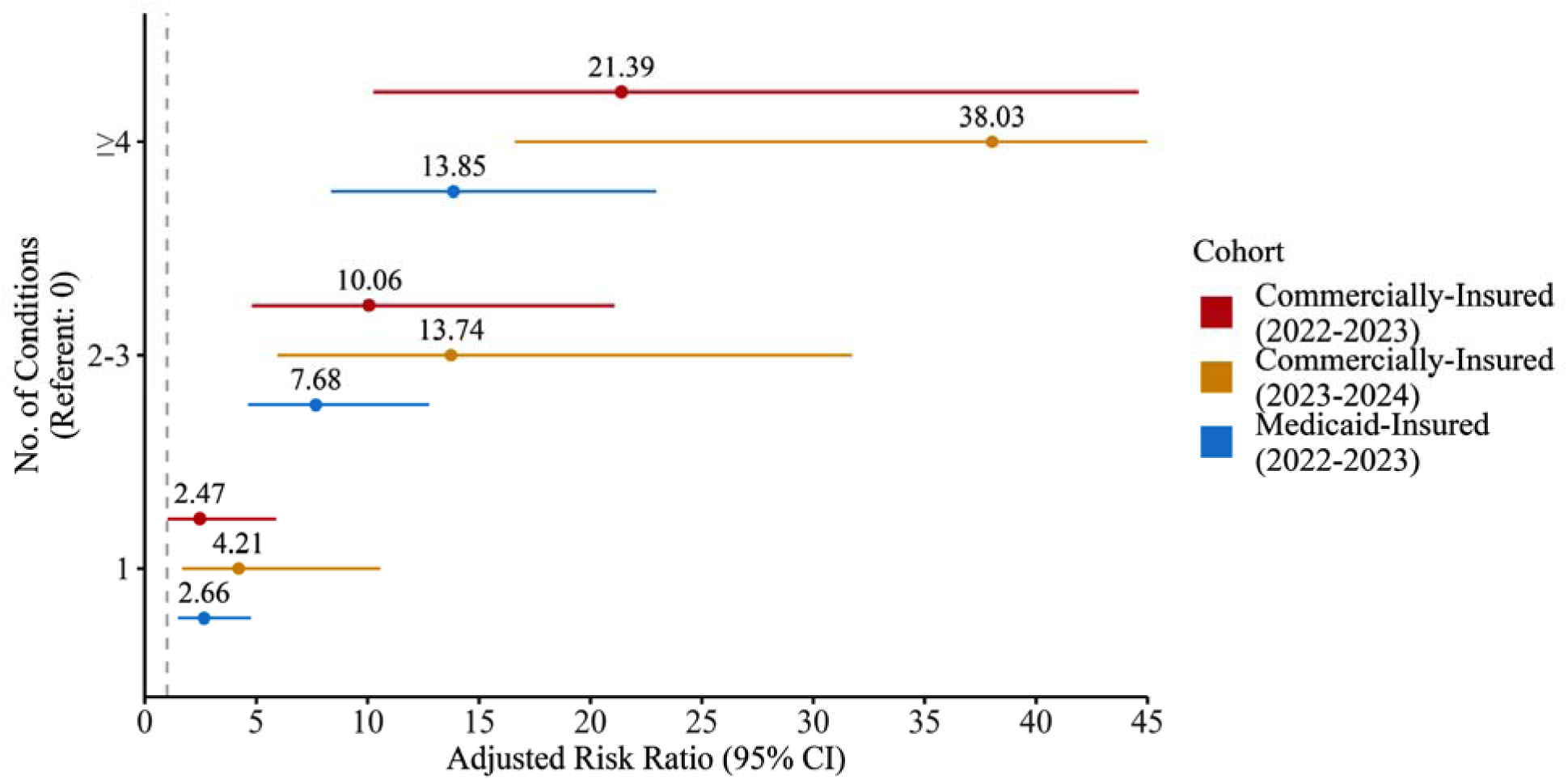
Adjusted Risk Ratios for Severe RSV Outcomes by Number of Underlying Chronic Conditions in Commercially- (2022–2024) and Medicaid-Insured (2022–2023) Cohorts. Footnote: Estimates shown are from a single model adjusted for age and sex. Referent group = individuals with 0 chronic conditions. Abbreviations: aRR: Adjusted Risk Ratio; CI: Confidence Interval; RSV: Respiratory Syncytial Virus.

Analysis of individual chronic conditions showed that individuals with chronic respiratory disease were at the highest risk of severe RSV outcomes in both cohorts. In the commercially-insured cohort, aRRs were 3.78 (95% CI: 2.72–5.23) in the 2022–23 season and 6.39 (95% CI: 4.54–9.00) in 2023–24; in the Medicaid-insured cohort, the aRR was 5.24 (95% CI: 3.94–6.98). Cardiovascular disease was the next leading condition associated with increased risk of severe RSV outcomes, with aRRs of 2.21 (95% CI: 1.52–3.21) and 2.00 (95% CI: 1.44–2.79) in the commercially insured cohort, and 1.77 (95% CI: 1.44–2.17) in the Medicaid-insured cohort.

Other comorbidities associated with increased risk included obesity, neurologic conditions, blood disorders, and chronic liver disease (**Figure 4**).

**Figure 4:**
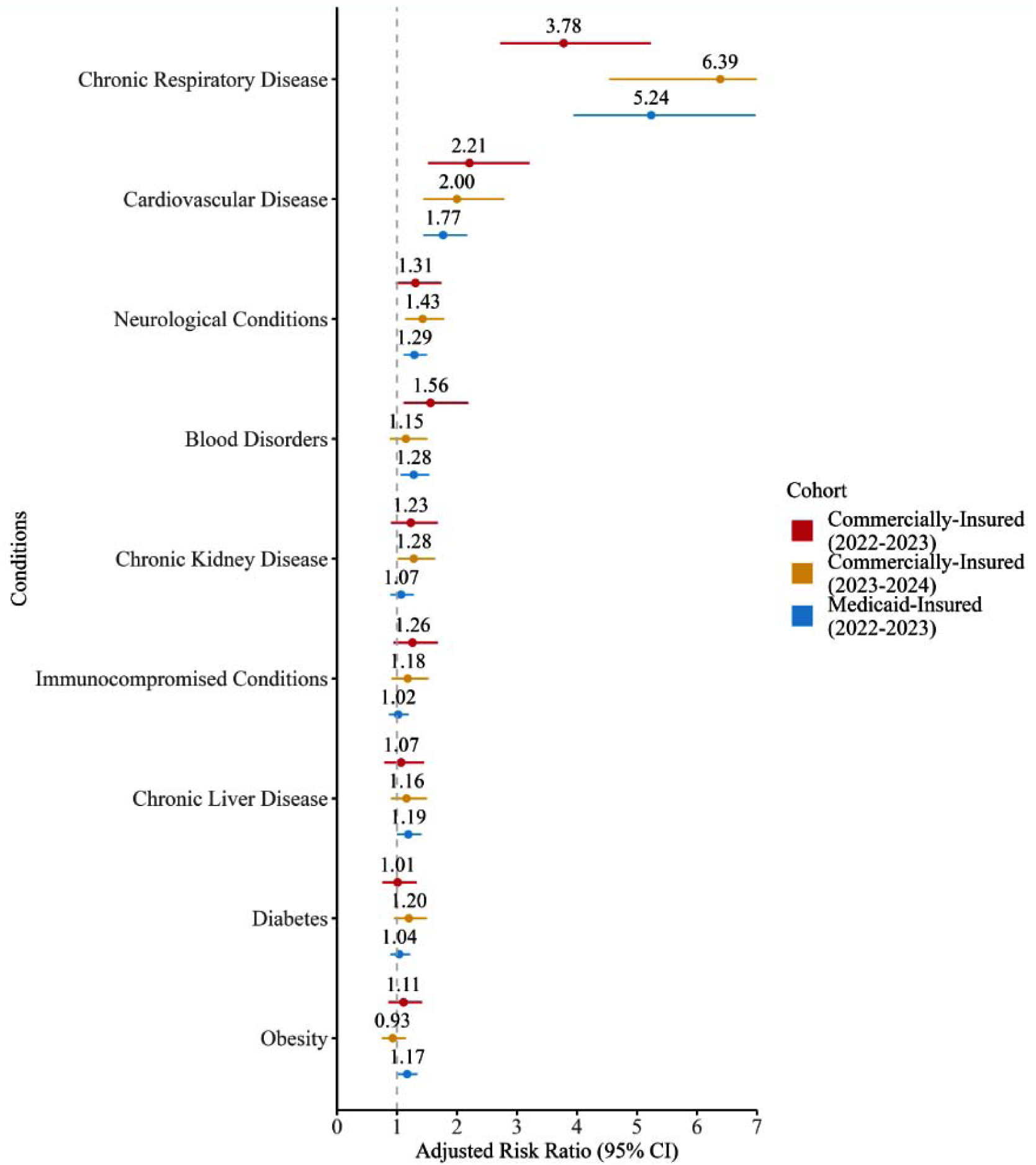
Adjusted Risk Ratios for Severe RSV Outcomes by Individual Underlying Chronic Conditions in Commercially- (2022–2024) and Medicaid-Insured (2022–2023) Cohorts. Footnote: Estimates shown are from a single model adjusted for age and sex. Abbreviations: aRR: Adjusted Risk Ratio; CI: Confidence Interval; RSV: Respiratory Syncytial Virus.

### Sensitivity Analyses

In sensitivity analyses restricted to individuals with RSV coded in the primary diagnosis position, the prevalence of chronic conditions was slightly lower compared with the main analysis; however, overall cohort characteristics remained consistent (**Supplementary Tables S7** and **S8**). When evaluating the associations between chronic conditions and severe RSV outcomes using narrower definitions to capture more severe forms of chronic diseases, the results were consistent with the main analysis, generally showing stronger measures of association (**Supplementary Table S9**).

Chronic conditions were associated with an increased risk of severe RSV outcomes across all adult age groups. In the Medicaid-insured cohort, individuals aged 18 years with at least one chronic condition had a 5.6-fold higher risk of severe RSV outcomes compared with those without chronic conditions (aRR = 5.62; 95% CI: 2.84–11.11). This risk progressively increased with age, reaching an aRR of 11.30 (95% CI: 3.61–35.32) at age 59 years. In the commercially-insured cohort, individuals aged 20 years with at least one chronic condition had a 2.9-fold higher risk of severe RSV outcomes in the 2022–23 season (aRR = 2.94; 95% CI: 0.88–9.79) and an 8.4-fold higher risk in 2023–24 (aRR = 8.36; 95% CI: 3.18–21.93). This risk increased with age, reaching an aRR of 24.42 (95% CI: 4.92–121.33) at age 50 years in 2022–23 and 21.99 (95% CI: 9.14–52.90) in 2023–24 (**Supplementary Table S10**).

## Discussion

This retrospective cohort study found that U.S. adults aged 18–59 years with medically attended RSV (MA-RSV) frequently had underlying chronic conditions. Approximately 75% had at least one chronic condition and over 25% had four or more. More than one in five adults with MA-RSV experienced severe outcomes, including hospitalization, ICU admission, and MV. The risk of severe outcomes substantially increased with the number of chronic conditions.

These findings highlight the vulnerability of a substantial portion of the working-age U.S. population. In our study, three in four adults aged 18–59 years with medically attended RSV had at least one chronic condition, and a quarter had four or more. This is substantially higher than the prevalence of chronic conditions in the general U.S. adult population. Data from the 2018 National Health Interview Survey (NHIS), part of the National Health and Nutrition Examination Survey (NHANES) program, show that approximately 51.8% of U.S. adults aged 18 and older had at least one diagnosed chronic condition [20]. Cardiovascular disease, chronic respiratory conditions, immunocompromised status, obesity, and diabetes were shown to increase the risk of severe RSV outcomes. More than one-third of adults in this age group are estimated to have at least one of these risk conditions that are associated with severe RSV outcomes, representing over 60 million individuals [21]. These data underscore the size of a high-risk population that may benefit from enhanced prevention efforts.

Across both commercially and publicly insured cohorts, severe RSV outcomes (hospitalization, ICU admission, oxygen support, and MV) were common, and their frequency increased with age and underlying multimorbidity. However, even among 18-year-olds (Medicaid cohort), having any chronic conditions conferred a 5-fold higher risk than healthy individuals. The highest risks were observed among individuals with chronic respiratory disease and cardiovascular disease, consistent with available literature on RSV and other respiratory pathogens [1,17,22,23]. Notably, individuals with four or more chronic conditions experienced a 14- to 38-fold higher risk of severe outcomes than healthy individuals.

Younger adults with chronic conditions may have a similar likelihood of developing severe RSV outcomes as older adults. Our results support existing literature showing that comorbidity, in addition to age, is a key driver of RSV-related morbidity [24]. Despite the high burden of chronic conditions among adults aged 18–59, current RSV vaccination recommendations from ACIP target adults aged ≥75 and those aged 60–74 years with specific risk conditions. This leaves a significant portion of high-risk younger adults without routine protection against RSV, despite immunogenicity data suggesting that adults under 60 years with chronic conditions respond well to RSV vaccination [25]. Our findings reinforce the need to broaden RSV prevention strategies beyond age-based thresholds to include younger adults with comorbidities.

Differences in healthcare utilization at the time of RSV diagnosis were observed between commercially insured and Medicaid-insured individuals. Medicaid enrollees were more often diagnosed during emergency or inpatient encounters, while commercial enrollees were typically diagnosed in outpatient settings. These patterns likely reflect disparities in healthcare access, provider availability, and care-seeking behavior—factors that should be considered when designing equitable RSV prevention and diagnostic strategies [26,27].

This study has several limitations. First, RSV diagnoses and comorbidities were identified via ICD-10-CM codes in claims data, which may be prone to misclassification. This limitation is particularly relevant given the historically limited use of RSV testing in adults, which could reduce the sensitivity of diagnostic coding and contribute to selection bias toward more severe cases. Second, mortality data were unavailable for the Medicaid cohort and only available as month of death for the commercial cohort, limiting our ability to assess deaths. Third, we used healthcare utilization (e.g., hospitalization, ICU admission) as proxies for disease severity, which may reflect access or utilization differences rather than true clinical severity. Objective clinical data (e.g., oxygen saturation, lab-confirmed severity scores) were not available. Fourth, the interrelated nature of comorbidities also complicates estimation of independent associations. To address this, we analyzed both individual conditions and the number of chronic conditions as risk indicators to capture the burden of multimorbidity. Lastly, because the data sources used in this study do not capture uninsured individuals, traditional Medicare beneficiaries, or patients with end-stage renal disease, these populations were not represented in the study cohort. This may limit the generalizability of our findings to the broader U.S. adult population.

This study demonstrates that chronic conditions are highly prevalent among U.S. adults aged 18– 59 years who seek medical care for RSV, and their presence significantly increases the risk of severe RSV outcomes, including hospitalization, ICU admission, mechanical ventilation, and death. The risk of severe illness rises sharply with the number of chronic conditions. These findings highlight the need for more inclusive and risk-based prevention strategies, such as vaccination, early diagnosis, and clinical guidance, to reduce RSV-related morbidity and healthcare burden in this vulnerable and often overlooked population.

## Data Availability

All data produced in the present study are available upon reasonable request to the authors

## Abbreviation list

aRR: Adjusted Risk Ratio
CAD: Coronary Artery Disease
CDC: Centers for Disease Control and Prevention
CDM: Optum’s de-identified Clinformatics® Data Mart Database
CI: Confidence Interval
COPD: Chronic Obstructive Pulmonary Disease
ED: Emergency Department
HF: Heart Failure
HCRU: Healthcare Resource Utilization
ICD-10-CM: International Classification of Diseases, Tenth Revision, Clinical Modification
ICU: Intensive Care Unit
IQR: Interquartile Range
LRTI: Lower Respiratory Tract Infection
MA-RSV: Medically Attended Respiratory Syncytial Virus
MDCD: Merative™ MarketScan® Multi-State Medicaid Database
MV: Mechanical Ventilation
RSV: Respiratory Syncytial Virus
RSV-NET: Respiratory Syncytial Virus Hospitalization Surveillance Network
SD: Standard Deviation
U.S.: United States

## Declarations

### Ethics approval and consent to participate

All data was fully deidentified and compliant with the Health Insurance Portability and Accountability Act (HIPAA) requirements. As this analysis involved secondary data without patient identifiers or direct contact, institutional review board (IRB) approval and informed consent were not required. The study followed the Strengthening the Reporting of Observational Studies in Epidemiology (STROBE) guidelines for observational research.

### Consent for publication

Not applicable

### Availability of data and materials

This study used de-identified, licensed administrative claims data from Optum Clinformatics® Data Mart (CDM) and Merative™ MarketScan® Multi-State Medicaid Database (MDCD).

These data are not publicly available due to licensing restrictions but may be obtained from the respective data vendors (Optum and Merative) upon reasonable request and with appropriate data use agreements. The authors do not have permission to share the datasets. All data used were fully de-identified following HIPAA requirements.

### Authors’ contributions

EW and ZZ conceived the study. DKN, LL, JS, NR, MW, IL, MJ, GG, EW, and ZZ developed the study protocol. LL, JS, MW, IL, and MJ conducted the data extraction and statistical analyses. JS and IL created the data visualizations. DKN, NR, and MW drafted the initial manuscript and coordinated revisions. All authors contributed to interpretation of the findings. NR and GG managed the project. EW and ZZ supervised the study. All authors reviewed and revised the manuscript and approved the final version.

### Authors’ information

Aetion Iberia, S.L., Barcelona, Spain Daniel K. Nomah

Moderna Tx, Cambridge, MA, USA

Linwei Li, Meng Wang, Miao Jiang, Genelle Goodhue, Beverly Francis, Sophia Monaghan, Claire Newbern, Catherine A. Panozzo, Elissa Wilker, Zhe Zheng

Aetion Inc., New York, NY, USA

John Shen, Neloufar Rahai, Isabella Lelis

