## Supplementary Materials for "Chronic conditions and risk of severe outcomes among adults aged 18-59 years with medically-attended RSV illness in the United States"

**Supplementary Material**

**Data Sources**

This study used deidentified administrative claims data from two large U.S. insurance databases: Optum® Clinformatics® Data Mart (CDM) and Merative™ MarketScan® Multi-State Medicaid Database (MDCD). Both databases contain comprehensive longitudinal data, including medical services, pharmacy prescriptions, and enrollment information. All data are deidentified and fully compliant with the Health Insurance Portability and Accountability Act (HIPAA). Because the study used secondary data without direct patient contact, institutional review board (IRB) approval was not required.

Optum CDM comprises adjudicated claims data for commercially insured members from a large national managed care organization. It provides geographically diverse coverage across the U.S. and represents approximately 19% of the U.S. commercially insured population. Medical claims in Optum CDM include inpatient, outpatient (including emergency department [ED]), physician-administered services, and outpatient laboratory test results from major national labs. Claims are verified and adjudicated before deidentification, ensuring accuracy and completeness. The dataset allows for a comprehensive view of patients’ healthcare utilization during their eligibility periods.

Merative™ MarketScan® MDCD includes claims data for approximately 12 million Medicaid enrollees aged 18–59 across multiple U.S. states and multiple Medicaid payers, offering critical insight into healthcare access and burden among low-income populations. Medicaid enrollees are more likely to experience barriers to care and may present with more advanced illness or comorbidity burden, making this dataset essential for characterizing RSV risk in vulnerable groups.

For this analysis, the RSV season was defined as July 1 through June 30 of the following year. Optum CDM was used to evaluate two RSV seasons: July 1, 2022, to June 30, 2023, and July 1, 2023, to June 30, 2024. Merative™ MarketScan® MDCD data were available for the 2022–2023 season only due to data release timelines.

Analyses were conducted separately for each database using consistent definitions and methodology. The follow-up period for assessing severe RSV outcomes and healthcare resource use was 30 days after the MA-RSV index date.

**Figure S1: Study Design Schema**

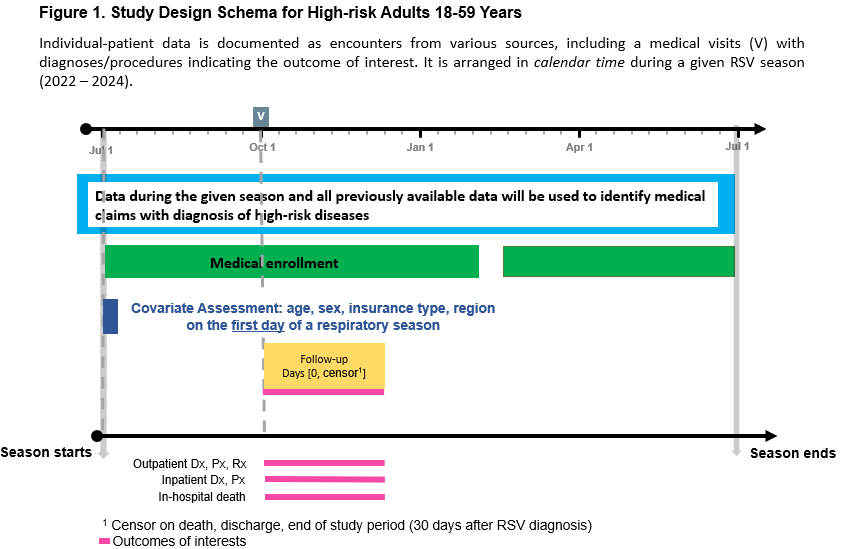

**Table S1: ICD-10-CM Codes Used to Define Medically attended RSV**

| **ICD-10-CM Code** | **Classification** | **Diagnosis Description** |
| --- | --- | --- |
| J21.0 | Medically attended RSV | Acute bronchiolitis due to respiratory syncytial virus |
| J12.1 | Medically attended RSV | Respiratory syncytial virus pneumonia |
| J20.5 | Medically attended RSV | Acute bronchitis due to respiratory syncytial virus |
| B97.4 | Medically attended RSV | Respiratory syncytial virus as the cause of diseases classified elsewhere |
| J12.9 | Unspecified LRTI | Viral pneumonia, unspecified |
| J17 | Unspecified LRTI | Pneumonia in diseases classified elsewhere |
| J18.0 | Unspecified LRTI | Bronchopneumonia, unspecified organism |
| J18.1 | Unspecified LRTI | Lobar pneumonia, unspecified organism |
| J18.2 | Unspecified LRTI | Hypostatic pneumonia, unspecified organism |
| J18.8 | Unspecified LRTI | Other pneumonia, unspecified organism |
| J18.9 | Unspecified LRTI | Pneumonia, unspecified organism |
| J20.9 | Unspecified LRTI | Acute bronchitis, unspecified |
| J21.9 | Unspecified LRTI | Acute bronchiolitis, unspecified |
| J22 | Unspecified LRTI | Unspecified acute lower respiratory infection |

**Table S2: Attrition Tables of Patients in the A) Commercially-Insured and B) Medicaid-Insured Cohorts**

A) **MA-RSV cohort**

|  | **2022-23 Season** | | **2023-24 Season** | |
| --- | --- | --- | --- | --- |
|  | **Number (%) of excluded** | **Number of individuals remaining** | **Number (%) of excluded** | **Number of individuals remaining** |
| Number of individuals in the dataset |  | 84,415,026 |  | 84,415,026 |
| Individuals meeting the inclusion criterion (MA-RSV diagnosis in the season of interest) |  | 31,536 |  | 33,637 |
| Excluded based on insufficient baseline enrollment | 15,062 (48%) | 16,474 | 14,424 (43%) | 19,213 |
| Excluded based on missing age | 0 (0%) | 16,474 | 0 (0%) | 19,213 |
| Excluded based on age, <18 years at index | 8,428 (51%) | 8,046 | 6,849 (36%) | 12,364 |
| Excluded based on age, >59 years at index | 6,611 (82%) | 1,435 | 10,473 (85%) | 1,891 |
| Excluded based on missing or unknown gender | 0 (0%) | 1,435 | 1 (<1%) | 1,890 |
| Individuals meeting all criteria |  | **1,435** |  | **1,890** |

**MA-RSV cohort with follow-up data**

|  | **2022-23 Season** | | **2023-24 Season** | |
| --- | --- | --- | --- | --- |
|  | **Number (%) of excluded** | **Number of individuals remaining** | **Number (%) of excluded** | **Number of individuals remaining** |
| Number of individuals in the dataset |  | 84,415,026 |  | 84,415,026 |
| Individuals meeting the inclusion criterion (MA-RSV diagnosis in the season of interest) |  | 31,536 |  | 33,637 |
| Excluded based on insufficient baseline enrollment¹ | 15,064 (48%) | 16,472 | 14,425 (43%) | 19,212 |
| Excluded based on insufficient post-index enrollment | 493 (3%) | 15,979 | 625 (3%) | 18,587 |
| Excluded based on missing age | 0 (0%) | 15,979 | 0 (0%) | 18,587 |
| Excluded based on age, <18 years at index | 8,171 (51%) | 7,808 | 6,564 (35%) | 12,023 |
| Excluded based on age, >59 years at index | 6,433 (82%) | 1,375 | 10,197 (85%) | 1,826 |
| Excluded based on RSV vaccine off-label use for individuals under 60 years (-365, 30) | 0 (0%) | 1,375 | 2 (<1%) | 1,824 |
| Excluded based on missing or unknown gender | 0 (0%) | 1,375 | 1 (<1%) | 1,823 |
| Individuals meeting all criteria |  | **1,375** |  | **1,823** |

*Footnote: Abbreviations - MA-RSV: Medically attended Respiratory Syncytial Virus*

*¹ Note: Patients excluded on their first medically attended RSV diagnosis are reassessed for inclusion based on subsequent diagnoses. As a result, attrition numbers may slightly differ from those in the Primary Objective 1 cohort, reflecting exclusions during later diagnoses.*

B) **MA-RSV cohort**

|  | **Number (%) of excluded** | **Number of individuals remaining** |
| --- | --- | --- |
| Number of individuals in the dataset |  | 86,082,087 |
| Individuals meeting the inclusion criterion (MA-RSV diagnosis in the season of interest) | -85,979,185 (99.9%) | 102,902 |
| Excluded based on insufficient baseline enrollment | -67,269 (65.4%) | 35,633 |
| Excluded based on missing age | -89 (0.2%) | 35,544 |
| Excluded based on Age <18 | -32,421 (91.2%) | 3,123 |
| Excluded based on Age >59 | -1,058 (33.9%) | 2,065 |
| Individuals meeting all criteria |  | **2,065** |

**MA-RSV cohort with follow-up data**

|  | **Number (%) of excluded** | **Number of individuals remaining** |
| --- | --- | --- |
| Number of individuals in the dataset |  | 86,082,087 |
| Individuals meeting the inclusion criterion (MA-RSV diagnosis in the season of interest) | -85,979,185 (99.9%) | 102,902 |
| Excluded based on insufficient baseline enrollment | -67,269 (65.4%) | 35,633 |
| Excluded based on missing age | -89 (0.2%) | 35,544 |
| Excluded based on Age <18 | -32,421 (91.2%) | 3,123 |
| Excluded based on Age >59 | -1,058 (33.9%) | 2,065 |
| Excluded insufficient post-enrollment | -111 (5.4%) | 1,954 |
| Excluded based on RSV vaccine use in the baseline or study period | 0 (0.0%) | 1,954 |
| Individuals meeting all criteria |  | **1,954** |

**Table S3: Code Definitions for Chronic Conditions Included in the Study (Broad Definitions)**

| **Condition** | **Code type** | **Codes** |
| --- | --- | --- |
| Cardiovascular (Others) | ICD-10-CM | I46.2, I46.8, I46.9, I48.0, I48.1, I48.11, I48.19, I48.2, I48.20, I48.21, I48.3, I48.4, I48.91, I48.92, I49.01, I49.02, I49.1, I49.2, I49.3, I49.40, I49.49, I49.5, I49.8, I49.9, I50.1, I50.20, I50.21, I50.22, I50.23, I50.30, I50.31, I50.32, I50.33, I50.40, I50.41, I50.42, I50.43, I50.810, I50.811, I50.812, I50.813, I50.814, I50.82, I50.83, I50.84, I50.89, I50.9, I11.0, I42.8, I42.9, I43, I27.0, I27.2, I27.20, I27.21, I27.23, I27.24, I27.29, I27.89, I27.9, I01, I05*, I06*, I07*, I08*, I09*, I10*, I11*, I15*, I51*, I20*, I23*, I25*, I26*, I27*, I28*, I30*, I31*, I32*, I33*, I34*, I35*, I36*, I37*, I38*, I39*, I40*, I41*, I42*, I44*, I45*, I46*, I48*, I52*, I70*, I71*, I72*, I73*, I74*, I75*, I76*, I77*, I5A, Q20*, Q21*, Q22*, Q23*, Q24*, Q25*, Q26*, Q27*, Q28*, A52.0, B37.6, I09.0, I09.1, I09.2, I09.81, I09.89, I09.9, I13.0, I13.2, I25.5, I42.0, I42.5, I42.6, I42.7** |
| Heart failure | ICD-10-CM | I50, I50.2, I50.3, I50.4, I50.8, I50.81* |
| Coronary artery disease | ICD-10-CM | I20.0, I21, I21.01, I21.02, I21.09, I21.11, I21.19, I21.0, I21.1, I21.2, I21.21, I21.29, I21.3, I21.4, I21.9, I21.A1, I21.A9, I22, I22.0, I22.1, I22.2, I22.8, I22.9, I24*, I24.0, I24.1, I24.8, I24.81, I24.89, I24.9, I25.1, I25.10, I25.11, I25.110, I25.111, I25.112, I25.118, I25.119, I25.2, I25.8, I25.81, I25.810, I25.811, I25.812, I25.82, I25.83, I25.84, I25.85, I25.89, I25.9** |
| Asthma | ICD-10-CM | J45, J45.2, J45.20, J45.21, J45.22, J45.3, J45.30, J45.31, J45.32, J45.4, J45.40, J45.41, J45.42, J45.5, J45.50, J45.51, J45.52, J45.9, J45.90, J45.901, J45.902, J45.909, J45.99, J45.990, J45.991, J45.998 |
| Chronic liver disease | ICD-10-CM | B18*, B19*, I85*, I86.4, I98.2, K70.0, K70.1*, K70.2*, K70.3*, K70.4*, K70.9, K71.1*, K71.3, K71.4, K71.5*, K71.6, K71.7, K71.8, K71.9, K72.1*, K72.9*, K73*, K74*, K75.2, K75.3, K75.4, K75.8, K75.9, K76.0, K76.1, K76.2, K76.3, K76.4, K76.5, K76.6, K76.7, K76.8*, K76.9, Z94.4 |
| Chronic kidney disease | ICD-10-CM | I12*, I12.9, I13.1*, I13.2*, N03*, N03.2, N03.3, N03.4, N03.5, N03.6, N03.7, N04*, N05*, N05.2, N05.3, N05.4, N05.5, N05.6, N05.7, N18*, N19*, N25*, Z49.0*, Z49.1*, Z49.2*, Z49.3*, Z94.0, Z99.2 |
| Diabetes | ICD-10-CM | E10*, E11*, E13* |
| Neurological conditions | ICD-10-CM | G04, G05, G10*, G11*, G12*, G13*, G14*, G20*, G21*, G23*, G30*, G31*, G32*, G35*, G36*, G37*, G45*, G46*, G70*, G71*, G72*, G73*, G80*, G81*, G82*, G83*, G91*, G93.1, G93.4, G93.40, G93.41, G93.49, G95*, I60*, I61*, I62*, I63*, I65*, I66*, I67*, I68*, I69*** |
| Blood disorders | ICD-10-CM | D55*, D56.0, D56.1, D56.2, D56.4, D56.5, D56.8, D56.9, D57.0*, D57.1, D57.2*, D57.4*, D57.8*, D58*, D59*, D60*, D61*, D64.0, D64.1, D64.2, D64.3, D64.4, D64.8*, D65*, D66*, D67*, D68* |
| Immunocompromised conditions | ICD-10-CM | C81*, C82*, C83*, C84*, C85*, C86*, C88*, C90*, C91*, C92*, C93*, C94*, C95*, C96*, D46*, D61.0*, D61.2, D61.9, D70.0, D71*, C00*, C01*, C02*, C03*, C04*, C05*, C06*, C07*, C08*, C09*, C10*, C11*, C12*, C13*, C14*, C15*, C16*, C17*, C18*, C19*, C20*, C21*, C22*, C23*, C24*, C25*, C26*, C27*, C28*, C29*, C30*, C31*, C32*, C33*, C34*, C35*, C36*, C37*, C38*, C39*, C40*, C41*, C42*, C43*, C44*, C45*, C46*, C47*, C48*, C49*, C50*, C51*, C52*, C53*, C54*, C55*, C56*, C57*, C58*, C59*, C60*, C61*, C62*, C63*, C64*, C65*, C66*, C67*, C68*, C69*, C70*, C71*, C72*, C73*, C74*, C75*, C76*, C77*, C78*, C79*, C7A*, C7B*, C80*, Z51.0, Z51.1*, C4A*, T86.0*, T86.1*, T86.2*, T86.3*, T86.4*, T86.5*, T86.81*, T86.85*, D47.Z1, Z48.2*, Z94*, Z98.85, D86*, E85.1, E85.2, E85.3, E85.4, E85.8*, E85.9, G35*, J67.9*, L40.54, L40.59, L93.0*, L93.2*, L94*, M05*, M06*, M07*, M08*, M30*, M31.3*, M31.5*, M32*, M33*, M34*, M35.3*, M35.8*, M35.9*, M46*, T78.40*, D27.9, D72.89, D80*, D81.0, D81.1, D81.2, D81.4, D81.5, D81.6, D81.7, D81.8*, D81.9, D82*, D83*, D84*, D87.89, D89.0, D89.1, D89.3, D89.4*, D89.8*, D89.9, K70.3*, K70.4*, K72*, K74.3, K74.4, K74.5, K74.6, N04*, R18.0, B20*, B21*, B22*, B23*, B24*, B97.35, O98.7*, Z21*, M45, M45.0, M45.1, M45.2, M45.3, M45.4, M45.5, M45.6, M45.7, M45.8, M45.9 |
| Chronic lung disease | ICD-10-CM | A15, A16, A19*, A31.0, B33.4, E84*, I27.8, I27.81, I27.82, I27.83, I27.89, I27.9, J40, J41, J41.0, J41.1, J41.8, J42, J43, J43.0, J43.1, J43.2, J43.8, J43.9, J44, J44.0, J44.1, J44.8, J44.81, J44.89, J44.9, J45, J45.2, J45.20, J45.21, J45.22, J45.3, J45.30, J45.31, J45.32, J45.4, J45.40, J45.41, J45.42, J45.5, J45.50, J45.51, J45.52, J45.9, J45.90, J45.901, J45.902, J45.909, J45.99, J45.990, J45.991, J45.998, J47, J47.0, J47.1, J47.9, J4A, J4A.0, J4A.8, J4A.9, J60, J61, J62, J62.0, J62.8, J63, J63.0, J63.1, J63.2, J63.3, J63.4, J63.5, J63.6, J64, J65, J66, J66.0, J66.1, J66.2, J66.8, J67, J67.0, J67.1, J67.2, J67.3, J67.4, J67.5, J67.6, J67.7, J67.8, J67.9, J68, J68.0, J68.1, J68.2, J68.3, J68.4, J68.8, J68.9, J69*, J70.0, J70.1, J70.3, J70.4, J70.5, J70.8, J70.9, J81.1, J82.81, J82.83, J82.89, J84*, J85*, J86*, J90*, J91*, J92*, J93*, J94*, J95*, J96*, J98*, J99*, Q33*** |
| Obesity | ICD-10-CM | E66, E66.0, E66.01, E66.09, E66.1, E66.8, E66.9, Z68.3, Z68.30, Z68.31, Z68.32, Z68.33, Z68.34, Z68.35, Z68.36, Z68.37, Z68.38, Z68.39, Z68.4, Z68.41, Z68.42, Z68.43, Z68.44, Z68.45 |

**Table S4: Codes Used to Identify Key Study Outcomes**

| **Condition** | **Code type** | **Codes** |
| --- | --- | --- |
| Oxygen support | HCPCS/CPT | E0442, E0441, E0439, E0440, E0424, E0425, E0444, E0443, E0434, E0433, E0435, E0431, E0430, A4620, A4608, 31730, 31719, 0061U, A4623, A4624, E0446, A4575, E0487, A4627, E0470, E0472, E0471, E1353, E0447, K0742, E1392, K0671, K0738, K0741, E1390, E1401, E1402, E1403, E1404, E1400, E1385, E1380, E1379, E1378, E1377, E1384, E1383, E1382, E1381, E1391, E1406, E1405, E1354, E1352, E1358, E1356, E1357, E0485, E0486, 4030F, 94651, 94652, 94650, E0467, E0466, E0465, A0422, K0534, E0470, K0532, K0533, E0472, E0471, E0452, K0194, G8854, 94660, K0193, E0601, A4604, A7037, K0187, G8845, G8852, G8850, G8847, G8853, A7044, G8851, G8855, A7034, K0183, E0561, K0268, E0562, K0531, A7035, K0185, A7030, A7039, K0189, A7038, K0188, G8849, A7036, K0186, A4615, G8569, G8164, 31500, 91000, 89100, 89105, 43756, 43757, A0396, 94657, 94004, 94002, 94003, 94656, 4168F, 99440, A4483, E0481, 99504, 94662, 33989, 33988, 36822, 33957, 33958, 33959, 33962, 33963, 33964, 33965, 33966, 33969, 33984, 33985, 33986, 33951, 33952, 33953, 33954, 33955, 33956, 33946, 33947, 33948, 33949, 33987 |
|  | ICD-10-CM | Z99.81, Z99.11, Z99.1, Z92.81 |
|  | ICD-10-PCS | 5A09357, 5A09457, 5A09557, 5A09358, 5A09458, 5A09558, 09HN7BZ, 09HN8BZ, 0BH13EZ, 0BH17EZ, 0BH18EZ, 0CHY7BZ, 0CHY8BZ, 0DH57BZ, 0DH58BZ, 0WHQ73Z, 0WHQ7YZ, 5A09357, 5A09457, 5A09557, 5A1935Z, 5A1945Z, 5A1955Z, 5A15223, 5A1522F, 5A1522G, 5A1522H, 5A15A2F, 5A15A2G, 5A15A2H |
| Antibiotic Treatment | Generic name | paromomycin sulfate, sulfamethoxazole/trimethoprim, trimethoprim, nitrofurantoin macrocrystal, nitrofurantoin monohydrate/macrocrystals, nitrofurantoin, NYSTATIN/TETRACYCLINE HCL/HYDROCORTISONE/DIPHENHYDRAMINE HCL, thalidomide, dapsone, atovaquone/proguanil HCl, trifluridine/tipiracil HCl, metronidazole, metronidazole in sodium chloride, tinidazole, secnidazole, atovaquone, ethambutol HCl, rifampin/isoniazid, rifampin, rifabutin, isoniazid, pyrazinamide, cycloserine, capreomycin sulfate, rifapentine, ethionamide, rifampin/isoniazid/pyrazinamide, meropenem, ertapenem sodium, meropenem in 0.9 % sodium chloride, doripenem, meropenem/vaborbactam, gentamicin sulfate/sodium citrate, cephalexin, ceftazidime, cefprozil, ceftriaxone sodium, cefpodoxime proxetil, cefuroxime axetil, cefazolin sodium, cefuroxime sodium, cefadroxil, cefoxitin sodium, cefdinir, cefaclor, cefotetan disodium, cefazolin sodium/water for injection, sterile, cefepime HCl, cefotetan disodium in iso-osmotic dextrose, cefazolin sodium in 0.9 % sodium chloride, cefixime, cefepime HCl in dextrose 5 % in water, cefazolin sodium/dextrose 5 % in water, ceftriaxone sodium in iso-osmotic dextrose, cefotaxime sodium, cefazolin sodium/dextrose, iso-osmotic, cefiderocol sulfate tosylate, cefepime HCl in iso-osmotic dextrose, ceftazidime in dextrose 5 % and water, cefditoren pivoxil, cefoxitin sodium/dextrose, iso-osmotic, ceftazidime/avibactam sodium, ceftaroline fosamil acetate, valganciclovir HCl, ganciclovir sodium, ganciclovir, tobramycin, tobramycin in 0.225 % sodium chloride, tobramycin/nebulizer, aztreonam lysine, sulfasalazine, minocycline HCl, ciprofloxacin HCl, levofloxacin/dextrose 5 % in water, levofloxacin, ciprofloxacin lactate/dextrose 5 % in water, moxifloxacin HCl in sodium chloride, iso-osmotic, moxifloxacin HCl, ciprofloxacin, gemifloxacin mesylate, delafloxacin meglumine, moxifloxacin HCl in sodium acetate and sulfate, water, iso-osmotic, ofloxacin, ciprofloxacin/ciprofloxacin HCl, oritavancin diphosphate, dalbavancin HCl, telavancin HCl, clarithromycin, erythromycin base, fidaxomicin, erythromycin lactobionate, erythromycin ethylsuccinate, erythromycin stearate, mupirocin calcium, vancomycin HCL/balanced salt solution no.2/PF, cefuroxime sodium in 0.9 % sodium chloride/PF, moxifloxacin HCl in balanced salt solution no.2/PF, piperacillin sodium/tazobactam sodium, ampicillin sodium, amoxicillin/potassium clavulanate, penicillin V potassium, oxacillin sodium, ampicillin sodium/sulbactam sodium, ampicillin trihydrate, amoxicillin, nafcillin sodium, penicillin G potassium/dextrose-water, piperacillin and tazobactam in dextrose, iso-osmotic, dicloxacillin sodium, penicillin G potassium, penicillin G benzathine, oxacillin sodium in iso-osmotic dextrose, penicillin G benzathine/penicillin G procaine, penicillin G procaine, penicillin G sodium, nafcillin in dextrose, iso-osmotic, ticarcillin disodium/potassium clavulanate, lansoprazole/amoxicillin trihydrate/clarithromycin, omeprazole magnesium/amoxicillin trihydrate/rifabutin, omeprazole/clarithromycin/amoxicillin trihydrate, colloidal bismuth subcitrate/metronidazole/tetracycline HCl, doxycycline hyclate, diluent for lefamulin (10 mM citrate buffered 0.9 % sodium chloride), colistin (as colistimethate sodium), polymyxin B sulfate, bacitracin, sulfadiazine, doxycycline monohydrate, tetracycline HCl, tigecycline, eravacycline di-hydrochloride, omadacycline tosylate, demeclocycline HCl, sarecycline HCl, doxycycline calcium, clindamycin phosphate, vancomycin HCl, clindamycin HCl, linezolid in dextrose 5 % in water, daptomycin, clindamycin palmitate HCl, vancomycin in 0.9 % sodium chloride, gentamicin sulfate, linezolid, clindamycin phosphate/dextrose 5 % in water, gentamicin sulfate in sodium chloride, iso-osmotic, tedizolid phosphate, aztreonam, vancomycin HCl in sterile water, vancomycin HCl in water for injection (PEG-400, NADA), vancomycin in 5 % dextrose in water, lincomycin HCl, clindamycin phosphate in 0.9 % sodium chloride, amikacin sulfate, fosfomycin tromethamine, rifamycin sodium, bacitracin zinc/polymyxin B sulfate, lefamulin acetate, tobramycin sulfate, linezolid in 0.9 % sodium chloride, amikacin sulfate liposomal with nebulizer accessories, gentamicin sulfate/PF, tobramycin sulfate/sodium chloride, neomycin sulfate/polymyxin B sulfate, pentamidine isethionate, neomycin sulfate, aztreonam/dextrose-water, quinupristin/dalfopristin, rifaximin, plazomicin sulfate, streptomycin sulfate, chloramphenicol sodium succinate, imipenem/cilastatin sodium/relebactam, imipenem/cilastatin sodium. |
|  | HCPCS/CPT | C9116, J1335, J0713, J0714, J0696, J0690, S0074, J0692, J0698, J0697, J0295, J0290, J2700, S0032, J2510, J2540, J0560, J0580, J0561, J0530, J0550, J0540, J0559, J0558, S0040, J0770, S0077, S0073, J2010, S0080, S0016, J0278, S0072, 80150, 82112, J7682, J7685, 80200, 84810, J3260, 80202, J3370, J1580, 80170, 84695, J0742, J0743. |
| Healthcare setting (MarketScan) - Emergency room/ Outpatient | Place of Serivice | 23 |
|  | Procedure Group | 111 |
|  | Revenue Code | 0450, 0451, 0452, 0456, 0459, 0981 |
|  | Service Sub-category | 10120, 10220, 10320, 10420, 10520, 12220, 20120, 20220, 21120, 21220, 22120, 22320, 30120, 30220, 30320, 30420, 30520, 30620, 31120, 31220, 31320, 31420, 31520, 31620. |
|  | Procedure code (CPT/ HCPC) | 99281, 99282, 99283, 99284, 99285, 99288. |
| Intensive care unit / inpatient | Revenue Code | 0200, 0201, 0202, 0203, 0204, 0206, 0207, 0208, 0209, 0210, 0211, 0212, 0213, 0214, 0219. |

Abbreviations: **ICD-10-CM**: International Classification of Diseases, Tenth Revision, Clinical Modification; **CPT**: Current Procedural Terminology; **HCPCS**: Healthcare Common Procedure Coding System; **NDC**: National Drug Code. *Note: Codes used to identify oxygen support include those for mechanical ventilation, which are* ***bolded*** *where applicable.*

**Table S5: Code Definitions for Chronic Conditions Included in the Study (Narrow Definitions)**

| **Condition** | **Code type** | **Codes** |
| --- | --- | --- |
| Cardiovascular (Others) | ICD-10-CM | I46.2, I46.8, I46.9, I48.0, I48.1, I48.11, I48.19, I48.2, I48.20, I48.21, I48.3, I48.4, I48.91, I48.92, I49.01, I49.02, I49.1, I49.2, I49.3, I49.40, I49.49, I49.5, I49.8, I49.9, I50.1, I50.20, I50.21, I50.22, I50.23, I50.30, I50.31, I50.32, I50.33, I50.40, I50.41, I50.42, I50.43, I50.810, I50.811, I50.812, I50.813, I50.814, I50.82, I50.83, I50.84, I50.89, I50.9, I11.0, I42.8, I42.9, I43, I27.0, I27.2, I27.20, I27.21, I27.23, I27.24, I27.29, I27.89, I27.9, I01, I05*, I06*, I07*, I08*, I09*, I10*, I11*, I15*, I51*, I20*, I23*, I25*, I26*, I27*, I28*, I30*, I31*, I32*, I33*, I34*, I35*, I36*, I37*, I38*, I39*, I40*, I41*, I42*, I44*, I45*, I46*, I48*, I52*, I70*, I71*, I72*, I73*, I74*, I75*, I76*, I77*, I5A, Q20*, Q21*, Q22*, Q23*, Q24*, Q25*, Q26*, Q27*, Q28*, A52.0, B37.6, I09.0, I09.1, I09.2, I09.81, I09.89, I09.9, I13.0, I13.2, I25.5, I42.0, I42.5, I42.6, I42.7** |
| Heart failure | ICD-10-CM | I50, I50.2, I50.3, I50.4, I50.8, I50.81* |
| Coronary artery disease | ICD-10-CM | I20.0, I21, I21.01, I21.02, I21.09, I21.11, I21.19, I21.0, I21.1, I21.2, I21.21, I21.29, I21.3, I21.4, I21.9, I21.A1, I21.A9, I22, I22.0, I22.1, I22.2, I22.8, I22.9, I24*, I24.0, I24.1, I24.8, I24.81, I24.89, I24.9, I25.1, I25.10, I25.11, I25.110, I25.111, I25.112, I25.118, I25.119, I25.2, I25.8, I25.81, I25.810, I25.811, I25.812, I25.82, I25.83, I25.84, I25.85, I25.89, I25.9** |
| Asthma | ICD-10-CM | J45.5, J45.50, J45.51, J45.52, J45.9, J45.99, J45.990, J45.991, J45.998 |
| Chronic liver disease | ICD-10-CM | B18*, B19*, I85*, I86.4, K70.0, K70.1*, K70.2*, K70.3*, K70.4*, K70.9, K71.1*, K71.3, K71.4, K71.5*, K71.6, K71.7, K71.8, K71.9, K72.1*, K72.9*, K73*, K74*, K75.3, K75.4, K75.8, K75.9, K76.1, K76.2, K76.3, K76.4, K76.5, K76.6, K76.7, K76.8*, K76.9, Z94.4 |
| Chronic kidney disease | ICD-10-CM | I12.0, I13.11, I13.2*, N18*, Z49.0*, Z49.1*, Z49.2*, Z49.3*, Z99.2 |
| Diabetes | ICD-10-CM | E10*, E11*, E13* (excluding E10.9, E11.9, E13.9) |
| Neurological conditions | ICD-10-CM | G04, G12, G13*, G14*, G35*, I63*, I69*, G70*, G71*, G72*, G73*, G80*, G81*, G82*, G83*, G95*** |
| Blood disorders | ICD-10-CM | D55*, D56.0, D56.1, D56.2, D56.4, D56.5, D56.8, D56.9, D57.0*, D57.1, D57.2*, D57.4*, D57.8*, D58*, D59*, D60*, D61*, D64.0, D64.1, D64.2, D64.3, D64.4, D64.8*, D66*, D67* |
| Immunocompromised conditions | ICD-10-CM | C81*, C82*, C83*, C84*, C85*, C86*, C88*, C90*, C91*, C92*, C93*, C94*, C95*, C96*, D46*, D61.0*, D61.2, D61.9, D70.0, D71*, C00*, C01*, C02*, C03*, C04*, C05*, C06*, C07*, C08*, C09*, C10*, C11*, C12*, C13*, C14*, C15*, C16*, C17*, C18*, C19*, C20*, C21*, C22*, C23*, C24*, C25*, C26*, C27*, C28*, C29*, C30*, C31*, C32*, C33*, C34*, C35*, C36*, C37*, C38*, C39*, C40*, C41*, C42*, C43*, C44*, C45*, C46*, C47*, C48*, C49*, C50*, C51*, C52*, C53*, C54*, C55*, C56*, C57*, C58*, C59*, C60*, C61*, C62*, C63*, C64*, C65*, C66*, C67*, C68*, C69*, C70*, C71*, C72*, C73*, C74*, C75*, C76*, C77*, C78*, C79*, C7A*, C7B*, C80*, Z51.0, Z51.1*, C4A*, T86.0*, T86.1*, T86.2*, T86.3*, T86.4*, T86.5*, T86.81*, T86.85*, D47.Z1, Z48.2*, Z94*, Z98.85, D86*, E85.1, E85.2, E85.3, E85.4, E85.8*, E85.9, G35*, J67.9*, L40.54, L40.59, L93.0*, L93.2*, L94*, M05*, M06*, M07*, M08*, M30*, M31.3*, M31.5*, M32*, M33*, M34*, M35.3*, M35.8*, M35.9*, M46*, T78.40*, D27.9, D72.89, D80*, D81.0, D81.1, D81.2, D81.4, D81.5, D81.6, D81.7, D81.8*, D81.9, D82*, D83*, D84*, D87.89, D89.0, D89.1, D89.3, D89.4*, D89.8*, D89.9, K70.3*, K70.4*, K72*, K74.3, K74.4, K74.5, K74.6, N04*, R18.0, B20*, B21*, B22*, B23*, B24*, B97.35, O98.7*, Z21*, M45, M45.0, M45.1, M45.2, M45.3, M45.4, M45.5, M45.6, M45.7, M45.8, M45.9 |
| Chronic lung disease | ICD-10-CM | J40, J41, J41.0, J41.1, J41.8, J42, J45.2, J45.20, J45.21, J45.22, J45.3, J45.30, J45.31, J45.32, J45.4, J45.40, J45.41, J45.42, J45.90, J45.901, J45.902, J45.909, J47, J47.0, J47.1, J47.9, J68.2, J68.3, J68.4, J68.8, J68.9 |
| Obesity | ICD-10-CM | Z68.4, Z68.41, Z68.42, Z68.43, Z68.44, Z68.45 |

**Table S6: Severe RSV Outcomes and Healthcare Resource Utilization (HCRU) Within 30 Days Post-Index (First RSV Claim), Stratified by Presence of Chronic Conditions**

|  | **With chronic conditions** | **Without chronic conditions** | **With conditions (narrow definition)** | **Without conditions (narrow definition)** |
| --- | --- | --- | --- | --- |
|  | **Optum CDM 2022-23** | | | |
|  | N=1,022 | N=353 | N=832 | N=543 |
| RSV-associated Hospitalization; n (%) | 280 (27.4%) | 8 (2.3%) | 274 (32.9%) | 14 (2.6%) |
| Length of stay in hospital, median [IQR] | 5.00 [3.25, 8.00] | 3.50 [2.25, 4.00] | 5.00 [4.00, 8.00] | 3.50 [2.00, 4.00] |
| RSV-associated ICU admission; n (%) | 124 (12.1%) | 2 (0.6%) | 123 (14.8%) | 3 (0.6%) |
| Length of stay in ICU, median [IQR] | 7.00 [5.00, 12.75] | 2.50 [2.00, 3.00] | 7.00 [5.00, 12.00] | 3.00 [2.00, 13.00] |
| Oxygen Support; n (%) | 167 (16.3%) | 4 (1.1%) | 161 (19.4%) | 10 (1.8%) |
| Mechanical Ventilation; n (%) | 59 (5.8%) | 0 (0.0%) | 58 (7.0%) | 1 (0.2%) |
| Death; n (%) | 0 (0.0%) | 0 (0.0%) | 0 (0.0%) | 0 (0.0%) |
| Antibiotic treatment; n (%) | 332 (32.5%) | 86 (24.4%) | 264 (31.7%) | 154 (28.4%) |
|  | **Optum CDM 2023-24** | | | |
|  | N=1,404 | N=419 | N=1,153 | N=670 |
| RSV-associated Hospitalization; n (%) | 391 (27.8%) | 6 (1.4%) | 385 (33.4%) | 12 (1.8%) |
| Length of stay in hospital, median [IQR] | 5.00 [3.00, 8.00] | 3.00 [1.75, 3.25] | 5.00 [3.00, 8.00] | 3.00 [2.25, 4.00] |
| RSV-associated ICU admission; n (%) | 176 (12.5%) | 0 (0.0%) | 174 (15.1%) | 2 (0.3%) |
| Length of stay in ICU, median [IQR] | 6.50 [4.00, 12.00] | - | 6.50 [4.00, 12.00] | 6.50 [4.00, 9.00] |
| Oxygen Support; n (%) | 234 (16.7%) | 2 (0.5%) | 228 (19.8%) | 8 (1.2%) |
| Mechanical Ventilation; n (%) | 78 (5.6%) | 0 (0.0%) | 77 (6.7%) | 1 (0.1%) |
| Death; n (%) | 1 (0.1%) | 0 (0.0%) | 1 (0.1%) | 0 (0.0%) |
| Antibiotic treatment; n (%) | 434 (30.9%) | 98 (23.4%) | 368 (31.9%) | 164 (24.5%) |
|  | **Merative™ MarketScan® 2022-23** | | | |
|  | N=1,494 | N=460 | N=1,246 | N=708 |
| RSV-associated Hospitalization; n (%) | 464 (31.1%) | 15 (3.3%) | 453 (36.4%) | 26 (3.7%) |
| Length of stay in hospital, median [IQR] | 6.00 [4.00, 8.00] | 4.00 [2.00, 5.00] | 6.00 [4.00, 8.50] | 4.00 [2.00, 5.25] |
| RSV-associated ICU admission; n (%) | 212 (14.2%) | 2 (0.4%) | 210 (16.9%) | 4 (0.6%) |
| Length of stay in ICU, median [IQR] | 7.00 [5.00, 12.00] | 8.50 [5.00, 12.00] | 7.00 [5.00, 12.00] | 5.50 [3.50, 10.50] |
| Oxygen Support; n (%) | 289 (19.3%) | 13 (2.8%) | 278 (22.3%) | 24 (3.4%) |
| Mechanical Ventilation; n (%) | 89 (6.0%) | 2 (0.4%) | 88 (7.1%) | 3 (0.4%) |
| Death; n (%) | N/A | N/A | N/A | N/A |
| Antibiotic treatment; n (%) | 499 (33.4%) | 112 (24.3%) | 428 (34.3%) | 183 (25.8%) |

**Table S7: Sensitivity analysis: Baseline demographic and clinical characteristics of adults with RSV coded in the primary diagnosis position**

| **Characteristic** | **Optum CDM**  **(2022–23)**  **N=865** | **Optum CDM**  **(2023–24)**  **N=1,116** | **Merative™ MarketScan® MDCD**  **(2022–23)**  **N=1248** |
| --- | --- | --- | --- |
| **Age, in years** |  |  |  |
| Mean (SD) | 44.14 (11.98) | 45.32 (11.66) | 37.79 (12.82) |
| Median [IQR] | 47.00 [35.00, 55.00] | 48.50 [37.00, 55.00] | 37.00 [26.00, 49.00] |
| **Age groups, in years, n (%)** |  |  |  |
| 18–29 | 130 (15.0%) | 142 (12.7%) | 395 (31.7%) |
| 30–39 | 166 (19.2%) | 186 (16.7%) | 298 (23.9%) |
| 40–49 | 184 (21.3%) | 254 (22.8%) | 250 (20.0%) |
| 50–59 | 385 (44.5%) | 534 (47.8%) | 305 (24.4%) |
| **Gender, n (%)** |  |  |  |
| Female | 563 (65.1%) | 669 (59.9%) | 922 (73.9%) |
| Male | 302 (34.9%) | 447 (40.1%) | 326 (26.1%) |
| **Race/Ethnicity, n (%)¹** |  |  |  |
| White | 616 (71.2%) | 787 (70.5%) | 735 (58.9%) |
| Black | 0 (0.0%) | 0 (0.0%) | 255 (20.4%) |
| Hispanic | 249 (28.8%) | 327 (29.3%) | 144 (11.5%) |
| Other/Unknown | 0 (0.0%) | 0 (0.0%) | 61 (4.9%) |
| Missing | 0 (0.0%) | 2 (0.2%) | 53 (4.2%) |
| **Type of Healthcare Encounter at RSV Diagnosis** |  |  |  |
| Outpatient visit | 419 (48.4%) | 553 (49.6%) | 404 (32.4%) |
| ED visit | 316 (36.5%) | 410 (36.7%) | 735 (58.9%) |
| Hospitalization | 130 (15.0%) | 153 (13.7%) | 109 (8.7%) |
| **Underlying Conditions** |  |  |  |
| Any condition of interest (broad definition)² | 635 (73.4%) | 838 (75.1%) | 906 (72.6%) |
| Any condition of interest (narrow definition)³ | 516 (59.7%) | 679 (60.8%) | 719 (57.6%) |

Abbreviations - MA-RSV: Medically attended Respiratory Syncytial Virus; SD: Standard Deviation; IQR: Interquartile Range; ED visit: Emergency Department visit; Optum CDM: Optum Clinformatics® Data Mart; Merative™ MarketScan® MDCD.

¹ Hispanic ethnicity is no longer reported in Optum CDM (as of v0.9). Race in Optum CDM is in the process of changing from third-party imputed to a self-reported source.

² Broad definition of high risk for severe RSV, defined as diagnosis with one or more of the following underlying medical conditions in medical claims: Cardiovascular disease (e.g. heart failure, coronary artery disease, congenital heart disease), Obesity, Diabetes, Chronic kidney disease, Neurologic condition, Immunocompromised conditions (e.g. rheumatologic/autoimmune/inflammatory disease, hematological malignancy, solid malignancy, transplant, other intrinsic immune condition or immunodeficiency), Chronic respiratory disease (e.g. COPD, Asthma, interstitial lung disease, cystic fibrosis, and emphysema), Chronic liver disease, Blood disorders

³ Narrow definition of high risk for severe RSV, defined as diagnosis with one of the following underlying medical conditions in medical claims:

Chronic cardiovascular disease (e.g., heart failure, coronary artery disease, or congestive heart failure [excluding isolated hypertension]), Chronic lung or respiratory disease (e.g., COPD, emphysema, asthma, interstitial lung disease, or cystic fibrosis), End-stage renal disease or dependence on hemodialysis or other renal replacement therapy, Diabetes mellitus complicated by chronic kidney disease, neuropathy, retinopathy, or other end-organ damage, or requiring treatment with insulin or sodium-glucose cotransporter-2 (SGLT2) inhibitor, Neurological or neuromuscular conditions causing impaired airway clearance or respiratory muscle weakness (e.g., poststroke dysphagia, amyotrophic lateral sclerosis, or muscular dystrophy [excluding history of stroke without impaired airway clearance]), Chronic liver disease (e.g., cirrhosis), Chronic hematologic conditions (e.g., sickle cell disease or thalassemia), Severe obesity (body mass index ≥40 kg/m2), Moderate or severe immune compromise (e.g. active treatment for malignancies, SOT or islet transplant, hematopoietic cell transplant, primary immunodeficiency, autoimmune disease (e.g. lupus, psoriasis, multiple sclerosis))

**Table S8: Sensitivity Analysis: Prevalence of Chronic Conditions Among Adults With Medically Attended RSV Coded in the Primary Diagnosis Position**

|  | **Optum CDM**  **(2022–23)**  **N=865** | **95% CI** | **Optum CDM**  **(2023–24)**  **N=1,116** | **95% CI** | **Merative™ MarketScan® MDCD**  **(2022–23)**  **N=1248** | **95% CI** |
| --- | --- | --- | --- | --- | --- | --- |
| **Broad definition of underlying conditions¹** |  |  |  |  |  |  |
| **Cardiovascular disease, n (%)** | 410 (47.4%) | (44.1%, 50.7%) | 547 (49.0%) | (46.1%, 51.9%) | 489 (39.2%) | [36.5%, 41.9%] |
| Heart failure | 87 (10.1%) | (8.2%, 12.2%) | 134 (12.0%) | (10.2%, 14.0%) | 111 (8.9%) | [7.3%, 10.5%] |
| Coronary artery disease | 86 (9.9%) | (8.1%, 12.1%) | 143 (12.8%) | (11.0%, 14.9%) | 71 (5.7%) | [4.4%, 7.0%] |
| **Chronic respiratory disease, n (%)** | 363 (42.0%) | (38.7%, 45.3%) | 463 (41.5%) | (38.6%, 44.4%) | 562 (45.0%) | [42.3%, 47.8%] |
| Chronic obstructive pulmonary disease (COPD) | 128 (14.8%) | (12.6%, 17.3%) | 188 (16.8%) | (14.8%, 19.2%) | 201 (16.1%) | [14.1%, 18.1%] |
| Asthma | 216 (25.0%) | (22.2%, 28.0%) | 244 (21.9%) | (19.5%, 24.4%) | 301 (24.1%) | [21.7%, 26.5%] |
| **Obesity, n (%)** | 310 (35.8%) | (32.7%, 39.1%) | 393 (35.2%) | (32.5%, 38.1%) | 381 (30.5%) | [28.0%, 33.1%] |
| **Diabetes, n (%)** | 202 (23.4%) | (20.7%, 26.3%) | 263 (23.6%) | (21.2%, 26.1%) | 226 (18.1%) | [16.0%, 20.2%] |
| **Chronic kidney disease, n (%)** | 89 (10.3%) | (8.4%, 12.5%) | 135 (12.1%) | (10.3%, 14.1%) | 85 (6.8%) | [5.4%, 8.2%] |
| **Neurologic condition, n (%)** | 86 (9.9%) | (8.1%, 12.1%) | 119 (10.7%) | (9.0%, 12.6%) | 126 (10.1%) | [8.4%, 11.8%] |
| **Immunocompromised conditions, n (%)²** | 373 (43.1%) | (39.9%, 46.4%) | 512 (45.9%) | (43.0%, 48.8%) | 476 (38.1%) | [35.4%, 40.8%] |
| **Chronic liver disease, n (%)** | 76 (8.8%) | (7.1%, 10.9%) | 101 (9.1%) | (7.5%, 10.9%) | 90 (7.2%) | [5.8%, 8.6%] |
| **Blood disorders, n (%)** | 46 (5.3%) | (4.0%, 7.0%) | 77 (6.9%) | (5.6%, 8.5%) | 57 (4.6%) | [3.4%, 5.7%] |
| ***Number of underlying medical conditions*** |  |  |  |  |  |  |
| Mean (SD) | 2.26 (2.08) | - | 2.34 (2.14) | - | 2.00 (1.85) |  |
| Median [IQR] | 2.00 [0.00, 4.00] | - | 2.00 [1.00, 4.00] | - | 2.00 [0.00, 3.00] |  |
| ***Number of underlying medical condition categories, n (%)*** |  |  |  |  |  |  |
| 0 | 230 (26.6%) | (23.8%, 29.6%) | 278 (24.9%) | (22.5%, 27.5%) | 342 (27.4%) | [24.9%, 29.9%] |
| 1 | 165 (19.1%) | (16.6%, 21.8%) | 220 (19.7%) | (17.5%, 22.1%) | 255 (20.4%) | [18.2%, 22.7%] |
| 2-3 | 237 (27.4%) | (24.5%, 30.5%) | 300 (26.9%) | (24.4%, 29.6%) | 388 (31.1%) | [28.5%, 33.7%] |
| ≥4 | 233 (26.9%) | (24.1%, 30.0%) | 318 (28.5%) | (25.9%, 31.2%) | 263 (21.1%) | [18.8%, 23.3%] |

*Footnote: Abbreviations - MA-RSV: Medically attended Respiratory Syncytial Virus; SD: Standard Deviation; IQR: Interquartile Range*

*¹ Broad definition of high risk for severe RSV, defined as diagnosis with one or more of the following underlying medical conditions in medical claims:*

*Cardiovascular disease (e.g. heart failure, coronary artery disease, congenital heart disease)*

*Obesity*

*Diabetes*

*Chronic kidney disease*

*Neurologic condition*

*Immunocompromised conditions (e.g. rheumatologic/autoimmune/inflammatory disease, hematological malignancy, solid malignancy, transplant, other intrinsic immune condition or immunodeficiency)*

*Chronic respiratory disease (e.g. COPD, Asthma, interstitial lung disease, cystic fibrosis, and emphysema)*

*Chronic liver disease*

*Blood disorders*

*² Immunocompromised conditions include autoimmune and inflammatory diseases (e.g., rheumatologic/autoimmune/inflammatory disease, hematological malignancy, solid malignancy, transplant, other intrinsic immune condition or immunodeficiency)*

**Table S9: Sensitivity Analysis: Attributable Risk of Chronic Conditions (Narrow Definition) for Severe RSV Outcomes ¹ ²**

|  | **Optum 2022-23**  **N=1,375** | **Optum 2023-24**  **N=1,823** | **Merative™ MarketScan® 2022-23**  **N=1,954** |
| --- | --- | --- | --- |
|  | aRR (95% CI) | | |
| **Blood disorders** | 1.83 (1.15, 2.91) | 1.30 (0.94, 1.80) | 1.27 [1.05, 1.54] |
| **Cardiovascular disease** | 1.34 (1.01, 1.78) | 1.37 (1.07, 1.75) | 1.32 [1.14, 1.53] |
| **Chronic kidney disease** | 1.27 (0.90, 1.78) | 1.13 (0.86, 1.49) | 0.95 [0.79, 1.15] |
| **Chronic liver disease** | 0.84 (0.55, 1.29) | 1.19 (0.88, 1.60) | 1.09 [0.90, 1.32] |
| **Chronic respiratory disease** | 5.31 (3.89, 7.25) | 7.57 (5.57, 10.28) | 7.64 [5.90, 9.90] |
| **Diabetes** | 0.94 (0.70, 1.26) | 1.21 (0.96, 1.52) | 1.16 [0.99, 1.36] |
| **Immunocompromised conditions** | 1.24 (0.92, 1.66) | 1.16 (0.90, 1.50) | 1.07 [0.92, 1.23] |
| **Neurological conditions** | 1.35 (0.96, 1.90) | 1.22 (0.93, 1.59) | 1.07 [0.90, 1.27] |
| **Obesity** | 1.30 (0.98, 1.72) | 0.99 (0.77, 1.26) | 1.22 [1.05, 1.41] |

*Abbreviations - aRR: Adjusted Risk Ratio; 95% CI: 95% Confidence Interval; MA-RSV: Medically Attended Respiratory Syncytial Virus.*

*¹ Model: log(E[y]) = β0 + β1*Age group + β2*Sex + β3*CardiovascularDisease + β4*Obesity + β5*Diabetes + β6*ChronicKidneyDisease + β7*NeurologicCondition + β8*Immunocompromised + β9*ChronicRespiratoryDisease + β10*ChronicLiverDisease + β11*BloodDisorders*

*² Narrow definition of high risk for severe RSV, defined as diagnosis with one of the following underlying medical conditions in medical claims:*

*Chronic cardiovascular disease (e.g., heart failure, coronary artery disease, or congestive heart failure [excluding isolated hypertension])*

*Chronic lung or respiratory disease (e.g., COPD, emphysema, asthma, interstitial lung disease, or cystic fibrosis)*

*End-stage renal disease or dependence on hemodialysis or other renal replacement therapy*

*Diabetes mellitus complicated by chronic kidney disease, neuropathy, retinopathy, or other end-organ damage, or requiring treatment with insulin or sodium-glucose cotransporter-2 (SGLT2) inhibitor*

*Neurologic or neuromuscular conditions causing impaired airway clearance or respiratory muscle weakness (e.g., poststroke dysphagia, amyotrophic lateral sclerosis, or muscular dystrophy [excluding history of stroke without impaired airway clearance])*

*Chronic liver disease (e.g., cirrhosis)*

*Chronic hematologic conditions (e.g., sickle cell disease or thalassemia)*

*Severe obesity (body mass index ≥40 kg/m2)*

*Moderate or severe immune compromise (e.g. active treatment for malignancies, SOT or islet transplant, hematopoietic cell transplant, primary immunodeficiency, autoimmune disease (e.g. lupus, psoriasis, multiple sclerosis))*

**Table S10: Adjusted Risk Ratios (aRRs) for Severe RSV Outcomes by Age and Chronic Condition Status in A) Commercially and B) Medicaid-Insured Adults**

**A)**

|  | **Adjusted Risk Ratios (aRR)** | |
| --- | --- | --- |
|  | **Individuals with MA-RSV (2022-23 Season)**  **N=1,375** | **Individuals with MA-RSV (2023-24 Season)**  **N=1,823** |
| **Adjusted Risk ratio at different ages** |  |  |
| ≥1 chronic conditions vs none at age 20 | 2.94 (0.88, 9.79) | 8.36 (3.18, 21.93) |
| ≥1 chronic conditions vs none at age 30 | 5.95 (2.83, 12.51) | 11.54 (4.96, 26.85) |
| ≥1 chronic conditions vs none at age 40 | 12.05 (4.60, 31.56) | 15.93 (7.07, 35.89) |
| ≥1 chronic conditions vs none at age 50 | 24.42 (4.92, 121.33) | 21.99 (9.14, 52.90) |

**B)**

| **Adjusted Risk ratio at different ages** | **Individuals with MA-RSV Medicaid cohort (2022-23 season)**  **N=1,954**  **aRR (95% CI)** |
| --- | --- |
| ≥1 chronic conditions vs none at age 18 | 5.62 [2.84, 11.11] |
| ≥1 chronic conditions vs none at age 20 | 5.81 [3.09, 10.96] |
| ≥1 chronic conditions vs none at age 30 | 6.89 [4.19, 11.33] |
| ≥1 chronic conditions vs none at age 40 | 8.17 [4.49, 14.87] |
| ≥1 chronic conditions vs none at age 50 | 9.69 [4.11, 22.83] |
| ≥1 chronic conditions vs none at age 59 | 11.30 [3.61, 35.32] |

*Footnote: Abbreviations - aRR: Adjusted Risk Ratio; 95% CI: 95% Confidence Interval.*

*Model: log(E[y]) = β0 + β1*age (continuous) + β2*Sex + β3*any of the underlying medical conditions + β4 age*any of the underlying medical conditions*
